# Drivers of adaptive evolution during chronic SARS-CoV-2 infections

**DOI:** 10.1101/2022.02.17.22270829

**Authors:** Sheri Harari, Maayan Tahor, Natalie Rutsinsky, Suzy Meijer, Danielle Miller, Oryan Henig, Ora Halutz, Katia Levytskyi, Ronen Ben-Ami, Amos Adler, Yael Paran, Adi Stern

## Abstract

In some immunocompromised patients with chronic SARS-CoV-2 infection, dramatic adaptive evolution occurs, with substitutions reminiscent of those in variants of concern (VOCs). Here, we searched for drivers of VOC-like emergence by consolidating sequencing results from a set of twenty-seven chronic infections. Most substitutions in this set reflected lineage-defining VOC mutations, yet a subset of mutations associated with successful global transmission was absent from chronic infections. The emergence of these mutations might dictate when variants from chronic infections can dramatically spread onwards. Next, we tested the ability to predict antibody-evasion mutations from patient- and viral-specific features, and found that viral rebound is strongly associated with the emergence of antibody-evasion. We found evidence for dynamic polymorphic viral populations in most patients, suggesting that a compromised immune system selects for antibody-evasion in particular niches in a patient’s body. We suggest that a trade-off exists between antibody-evasion and transmissibility that potentially constrains VOC emergence, and that monitoring chronic infections may be a means to predict future VOCs.

## Introduction

Severe Acute Respiratory Syndrome Coronavirus 2 (SARS-CoV-2) infections normally resolve clinically within a few days, and RNA shedding lasts a few days to a few weeks ^1^. However, more and more case reports are accumulating that document chronic infections spanning weeks to many months of infection ^2,3^. Notably, chronic infection should not be confused with long-COVID, where infection is cleared rapidly, yet symptoms persist ^4^; in cases of chronic infections, replicative virus is detected for lengthy periods of time. To date, all cases of chronic SRAS-CoV-2 infection were associated with a severely immunosuppressed condition, ranging from primary immunodeficiencies, patients following solid organ transplants, acquired immunodeficiency syndrome (AIDS), patients undergoing therapy with immunosuppressive drugs, and hematologic cancers patients. Presumably, these immune system disorders prevent clearance of the virus as compared to patients with an intact immune system, and thus the virus thrives for lengthy periods of time ^5^.

Longitudinal sequencing of some cases of chronic infection has uncovered striking mutational patterns of evolution, and revealed that the rate of evolution is much higher than that observed along transmission chains of acutely infected individuals (e.g., ^6-8^). Indeed, several recent reports have found that the intra-host variation during acute infections is quite limited ^9-12^. Moreover, the transmission bottleneck size was inferred to be quite low in these studies, as evidenced by little to no shared genetic diversity in transmission pairs. Overall, this suggests that adaptive evolution may be limited during acute infections, and that most of the diversity observed during SARS-CoV-2 circulation is due to neutral mutations fixed during the small transmission bottleneck. The low genetic diversity observed in acute infections contrasts the high genetic diversity observed in rare cases of chronic infections.

Much interest has arisen in cases of chronic infection since the first emergence of variants of concern (VOCs). One major hypothesis that came up when the Alpha VOC was first discovered, and more recently with the rise of the Omicron variant, was that these variants may have evolved due to selection pressure created during chronic infections ^2,13^. More specifically, the hypothesis posited that strong selection for antibody-evasion mutations may occur in immunocompromised individuals with chronic infection who are treated with convalescent plasma (CP) or monoclonal antibodies (mAbs), collectively denoted here as antibody-based treatments (ABT).

However, there does not seem to be a consistent pattern that describes the evolution of SARS-CoV-2 across all chronic infections. Whereas some cases display dramatic evolution in the spike (s) protein (e.g., ^8^), in other chronic infections relatively limited evolution is observed (e.g., ^14^). Here, we set out to consolidate the evolutionary patterns found across chronic infections, by re-analyzing previous reports, and by sequencing a cohort of six patients with chronic SARS-CoV-2 infection patients at the Tel Aviv Sourasky Medical Center ^15,16^. We explore the shared and unique mutational patterns that emerge in different chronically infected patients, and compare them to those observed in global data reflecting transmission chains in acute individuals. We focus on inferring the conditions that lead to dramatic evolution and the potential for the creation of new VOCs.

## Results

### Different patterns of evolution detected in chronically infected patients

We begin by defining criteria for a chronic infection. In clinical settings, a chronic infection is often defined as one with both prolonged shedding of viral RNA, coupled with evidence for infectious virus (either through virus isolation in tissue culture, or via detection of sub-genomic RNA). However, when surveying various studies reporting chronic infection, a lack of standardization was noted. We hence expanded our focus to include patients displaying high-viral-load shedding for 20 or more days, while mining the literature for all such cases that were accompanied by longitudinal whole-genome sequencing of the virus (Methods). The criterion of 20 days was based on a meta-analysis of the duration of viral shedding (defined as a positive nasopharyngeal PCR test) across thousands of patients diagnosed up till June 2020, which revealed that mean duration of upper-respiratory tract shedding was around 17 days, with a 95% confidence interval ranging from 15.5 to 18 days ^17^. Of note, shedding of replication-competent virus lasted a lot less than twenty days. Moreover, estimates of viral shedding are different in some of the more recently detected SARS-CoV-2 variants such as Delta and Omicron ^18,19^, yet as described below, our analysis focused on variants that were found in earlier stages of the pandemic.

Our search yielded a total of 21 case reports, all of which were diagnosed during 2020 or early 2021, and all of which were infected with viruses belonging to lineages that predated the Alpha variant (Table S2). In addition, six patients adhering to the above criterion were detected in Tel Aviv Sourasky Medical Center (TASMC), and all available samples were sequenced (Methods). Five TASMC patients suffered from hematological cancers. The sixth patient suffered from an autoimmune disorder, and was treated with a high dose of steroids. The six TASMC patients were all diagnosed in late 2020 or early 2021, with four patients infected with a virus from pre-Alpha lineages, and two patients infected with a virus from the Alpha lineage (Table S2).

Overall, when observing the entire set of 27 chronically infected patients, we observed that all were immunosuppressed, due to one or more of the following: hematological cancer, anti-B cell treatment, high dosage steroid treatment, or very low CD4 counts (acquired immunodeficiency disease, AIDS). We observed very different evolutionary outcomes across the range of patients examined, from dramatic evolution and antibody-evasion observed in some patients, to relatively static evolution in others (Table 1, Tables S1-S2).

**Table 1.** Summary of all 27 patients with chronic SARS-CoV-2 infections. The last six patients were sequenced herein. See Tables S1,S2 for more details.

| Background condition | Anti-B cell background treatment<sup>a</sup> | Steroids treatment | Antibody-based COVID-19 treatment<sup>b</sup> | # days of infection<sup>c</sup> | Spike amino-acid replacements<sup>d</sup> | Ref. |
| --- | --- | --- | --- | --- | --- | --- |
| Chronic lymphocytic leukemia (CLL) | None | None | CP | 105 | None | 20 |
| Lymphoma | CD20 bispecific antibodies | Prednisone | CP | 131 | None | 14 |
| B cell lymphoma | CD20 bispecific antibodies | None | CP | 164 | V3G, Δ18-30, S50L, N87S, **Δ144-145**,A222V | 6 |
| Severe antiphospholipid syndrome | Rituximab | None | mAb | 150 | Y144F, **Δ141-143**, **E484K**, T478K, Q183H, **S494P**, **N501Y**, Q493K, Q183H, Q493H, P9L, **Y489H**, Δ12-18, **N440D**, A1020S, **Y144-**, **E484Q**, **F486I** | 8 |
| Kidney transplant | None | Prednisone | CP | 94 | **Δ141-144**, **E484K** | 21 |
| Renal disease | Rituximab | Prednisone | mAb | 20 | **E484K** | 22 |
| AIDS | None | None | mAb | 30 | **E484K**, Q954L | 22 |
| Follicular lymphoma | Obinutuzumab | None | mAb | 102 | **E484K** | 22 |
| Heart transplant | None | Prednisone | mAb, CP | 32 | **E484K** | 22 |
| CLL | None | None | mAb | 90 | G1219C, **E484K** | 22 |
| Kidney transplant | None | Prednisone | mAb | 26 | None | 22 |
| AIDS | None | None | None | 190 | **E484K**, A1078V, R190K, **K417T**, **F490S**, **D796H**, D427Y, **N501Y** | 23 |
| Marginal B cell lymphoma | Rituximab | None | CP | 102 | **Δ68-70**, **D796H**, Y200H, T240I, S13I, I68R, P330S, P812S | 7 |
| Acute lymphoblastic leukemia (ALL) | Inotuzumab | Dexamethasone | None | 79 | **Δ144**, S494P | 24 |
| AIDS, leukoencephalopathy | None | None | None | 91 | None | 25 |
| Heart transplanted, chronic kidney disease | None | Prednisone | None | 136 | None | 25 |
| ALL | low CD19 | Dexamethasone | CP | 144 | T95I, **Δ145**, **Δ141-144**, T2211I | 26 |
| ALL | low CD19 | None | None | 162 | S13I, L97M, A307V, **E484K**, **N440K** | 26 |
| Follicular lymphoma | Rituximab | Dexamethasone | HP | 189 | S172A | 27 |
| Follicular lymphoma | Rituximab | Dexamethasone | HP | 141 | D88E, I788V | 27 |
| Follicular lymphoma | Rituximab | Methyl-prednisolone | HP | 66 | None | 27 |
| CLL | Rituximab | None | mAb | 56 | **E484Q** | P1 |
| Follicular lymphoma | Obinutuzumab | None | None | 82 | None | P2 |
| Hodgkin's lymphoma | None | None | None | 68 | **G485R**, W258C | P3 |
| Autoimmune skin disease | None | Prednisone | None | 36 | None | P4 |
| ALL | Inotuzumab | None | CP | 89 | **E484K**, **F490L**, **Δ144** | P5 |
| CLL | None | Prednisone | CP | 37 | None | P6 |

<sup>a</sup> Yellow background indicates inferred B-cell depletion (Methods)
<sup>b</sup> Blue background indicates ABT during the period where virus was sequenced
<sup>c</sup> Shown is the maximal day-since-infection sequenced
<sup>d</sup> Green background indicates the presence of spike amino-acid substitutions, sorted by order of emergence, with antibody-evasion ones shown in bold (Methods, Table S1). Only mutations reaching a frequency higher than 80% at a given time point are shown

### Comparing patterns of evolution between chronic infections and global transmission chains

We searched for patterns of evolution across the total of twenty-seven patients with chronic infection, and compared this pattern with the pattern observed (a) under mostly neutral evolution, in the first approximately nine months of viral circulation ^28,29^, and (b) under presumable positive selection, which occurred in the lineages leading to the five currently defined VOCs (Alpha, Beta, Gamma, Delta, Omicron) (Fig. 1, Table S4). In each scenario, we searched for bins of mutations (along consecutive regions of 500 bases) enriched for mutations (p<0.05, binomial test, after correction for multiple testing; Methods).

**Figure 1.**
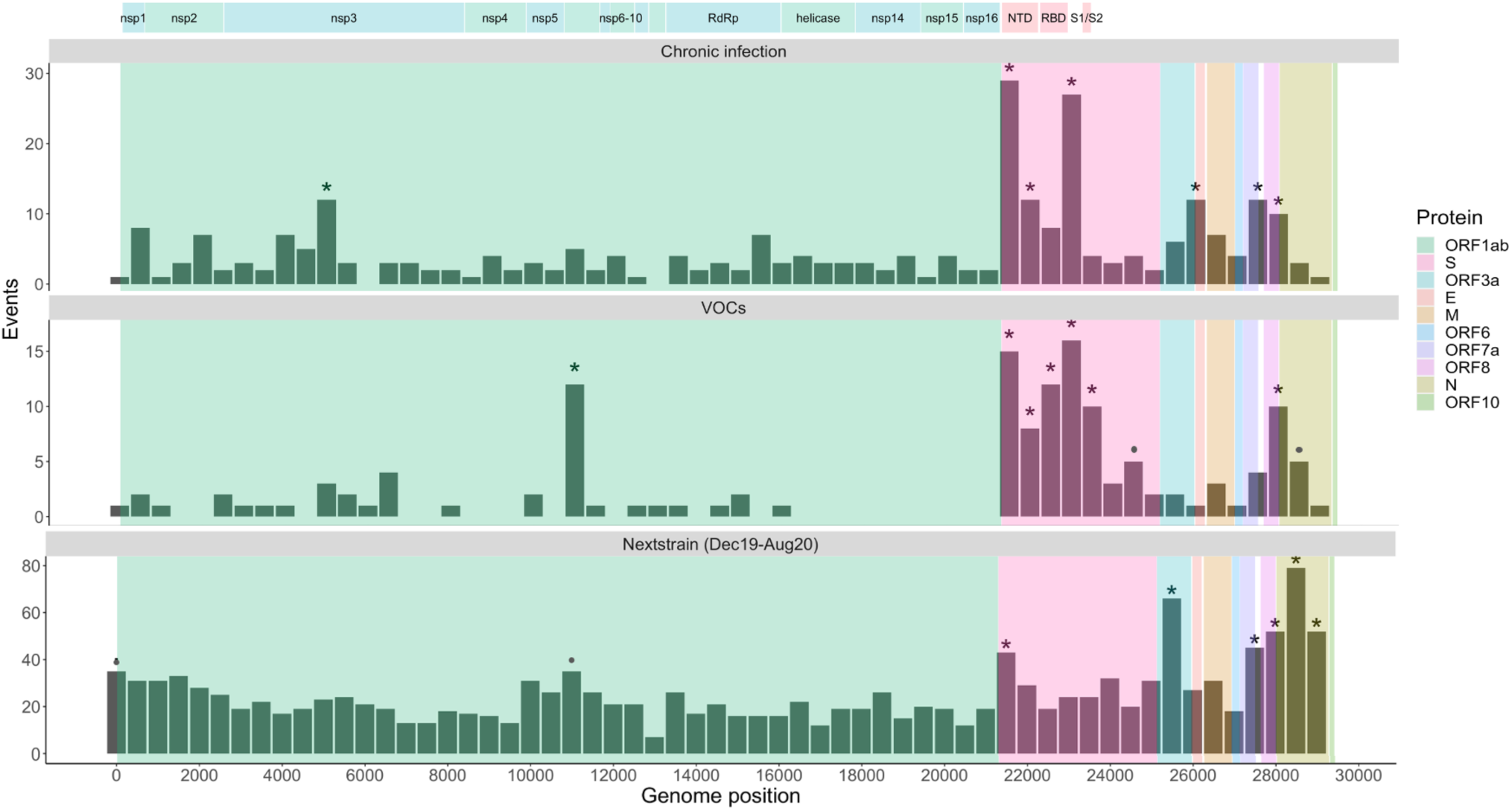
Number of substitutions observed along the SARS-CoV-2 genome, in bins of 500 nt. The upper panel displays substitutions observed at any time-point of the twenty-seven chronic infections. The middle panel displays lineage-defining mutations of the five currently recognized VOCs. The lower panel displays substitutions observed globally during the first nine months of the pandemic, mostly before the emergence of VOCs. Asterisks and dots mark bins enriched for more substitutions (P<0.05 and P<0.1, respectively, Methods). The genomic positions are based on the Wuhan-Hu-1 reference genome (GenBank ID NC_045512), and the banner on the top shows a breakdown of ORF1a/b into individual proteins, and domains for the spike protein (see main text).

**Figure 2.**
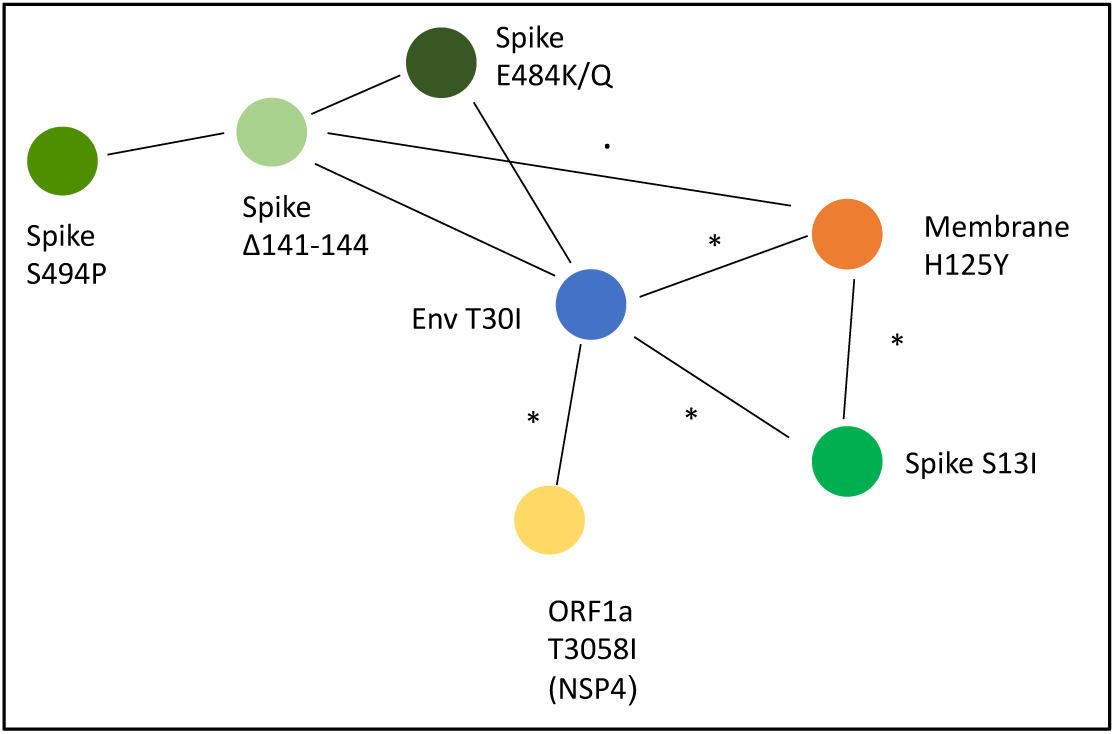
A network of co-occurring substitutions across patients with chronic SARS-CoV-2 infection. Each colored circle represents a locus, and an asterisk and dot represent P<0.05 and P<0.1, respectively.

During the first nine months of circulation, we noted that 65% of substitutions were non-synonymous, which is generally what we could expect under lack of both positive and purifying selection, and in line with reports suggesting incomplete purifying selection during the early stages of SARS-CoV-2 spread ^30^. During this time, we observed a relatively uniform distribution of substitutions across most of the genome, with some enrichment in ORF3a, ORF7, ORF8, and in N. This enrichment has been previously reported and may be due to more relaxed purifying selection in these regions, or higher mutation rates ^28^; adaptive evolution at these regions cannot be ruled out as well. We also noted some enrichment during the first nine months of the pandemic in the N-terminal domain (NTD) of the spike, which appeared to be mostly confined to the very first residues of the protein. These residues belong to the “NTD supersite” and are associated with immune evasion ^31,32^, which seems unlikely at this early stage of the pandemic, yet the NTD is also associated with structural rearrangements following ACE2 receptor binding^33^.

In general, the patterns obtained in chronic infections and in VOCs were very similar. The average proportion of non-synonymous substitutions was 78% and 82% in chronic infections and VOCs, respectively, generally suggestive of positive selection operating on both. The most striking similarity was observed in patterns along the Spike protein, and in particular at the regions which correspond to the NTD (genomic nucleotides 21,598 to 22,472) and the receptor binding domain (RBD) (genomic nucleotides 22,517 to 23,183). Several mutations at the RBD have been shown to enhance affinity to the ACE2 receptor and allow for better replication ^34,35^, whereas other mutations, both at RBD and NTD, are known to enhance antibody-evasion ^31,36,37^. Notably, chronic infections exhibited mutations both in the first NTD residues observed in the “neutral” pattern described above, but also in later positions. The most commonly observed substitutions in chronic infections were in the spike protein: E484K/Q and various deletions in the region spanning the NTD supersite, particularly amino-acids 140 through 145, all shown to confer antibody-evasion ^38^. Chronic infections shared the enrichment of ORF3a/ORF7a/ORF8 mutations with the “neutral” set, but lacked an enrichment across most of the N protein. Overall, it seems that mutations in chronic infections are predictive of those occurring in VOCs, as has been noted previously ^2^.

When focusing on the differences between VOCs and chronic infections, several intriguing differences emerged. First, four VOCs bear a three-amino acid deletion in the nsp6 gene (ORF1a: Δ3675-3677), an event not observed in our set of chronic infections. Next, in VOCs there is an enrichment in the region of the spike encompassing the S1/S2 boundary (positions 23,500-24,000 in Fig.1). This enrichment is primarily driven by S: P681H/R, a highly recurrent globally occurring mutation ^39^, surprisingly never observed in our chronic set. A recent analysis has analyzed the relationship between recurrent mutations and their success in driving clade expansion, i.e., onwards transmission ^40^. Notably, we observed that successful recurrent mutations were almost never observed in our chronic set, whereas non-successful recurrent mutations (S:E484K/Q and S:Δ144) were the ones most abundantly observed (Table 2). Overall, these results suggest that there may be a trade-off between antibody-evasion and transmissibility. This trade-off, if it exists, would not play a role in chronic infections, but would impact the ability of a variant created in a chronic infection to be transmitted onwards. Thus, only under specific conditions, a transmissible variant would emerge in chronic infections. Four of five VOCs independently acquired a mutation at or near the S1/S2 boundary (S:P681H/R or H655Y) suggesting that this may be a factor driving transmissibility. We note that Beta is an exception with no such mutations, yet this variant also displayed very limited global transmission.

**Table 2.** Recurrent mutations observed along the SARS-CoV-2 phylogeny. Clade success or lack of success is reported based on measurements of clade logistic growth, with + or – representing higher or lower than average growth, respectively ^40^. The last column marks the number of patients in the set of chronic infections herein where a substitution was observed.

| Protein | Mutation | Clade success | # times observed chronic |
| --- | --- | --- | --- |
| ORF1a | T3255I | + | 1 |
| ORF1a | S3675-,G3677-,F3678- | + | 0 |
| S | L18F | + | 0<sup>a</sup> |
| S | T95I | + | 1 |
| S | L452R | + | 0 |
| S | N501Y | + | 2 |
| S | P681R | + | 0 |
| N | P199L | + | 0 |
| ORF1a | L3606F | - | 1 |
| S | H69-,V70- | - | 1 |
| S | Y144- | - | 7<sup>b</sup> |
| S | E484K | - | 10<sup>c</sup> |
| S | P681H | - | 0 |
| N | S194L | - | 0 |
| N | T205I | - | 0 |
| N | M234I | - | 0 |

<sup>a</sup> Two deletion events were observed at this locus
<sup>b</sup> An additional two deletion events were observed at loci 141-143
<sup>c</sup> In three additional patients, E484Q was observed

We went on to examine co-occurring substitutions, defined as pairs of substitutions that appeared in two or more patients. We used a chi-square test to assess whether pairs of substitution occurred together more often than expected from their individual frequencies (Methods), as a measure of possible epistasis. Intriguingly, four pairs of substitutions across four different proteins emerged as significantly enriched, and formed a network of interactions: T30I in envelope, H125Y in the membrane glycoprotein, S13I in the spike protein, and T3058I in ORF1a (Fig. 4). This finding was intriguing on multiple fronts: first, envelope and membrane glycoprotein have generally remained very conserved throughout the entire pandemic, and specifically the two replacements found are at highly conserved sites (Table S1). The replacements in spike and ORF1a, on the other hand, have been observed a small number times in the global phylogeny. Notably, all the first three proteins form a part in the virion structure itself – however the functional meaning of this remains unclear. Other pairs of mutations found to co-occur were the three most common spike antibody-evasion mutations, yet these co-occurrences were not statistically significant. Larger cohorts of patients and further data will be required to determine the implications of these findings.

**Figure 3.**
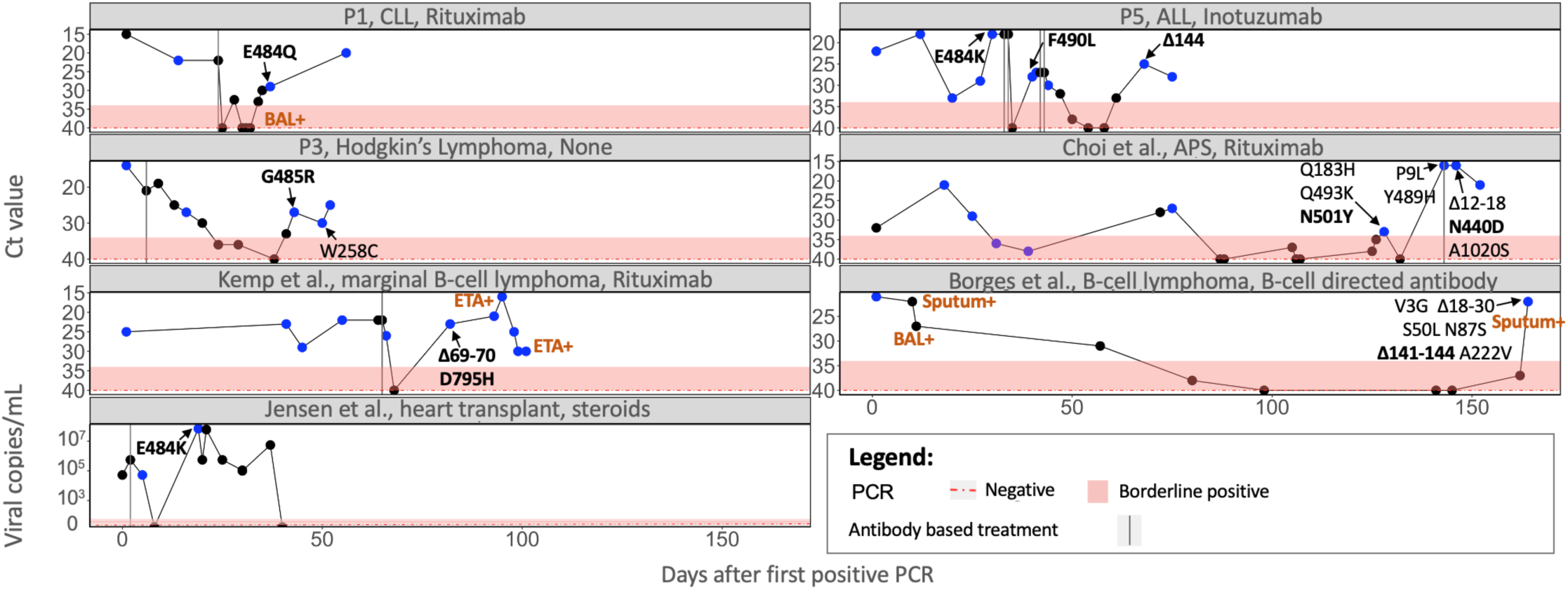
Viral rebound predicts mutations in the spike protein associated with immune evasion. Ct values are presented according to the day of infection (denoted as number of days post the first positive PCR test), with the dashed red horizontal line and shaded area representing a negative or borderline result, respectively. Blue dots represent samples that were sequenced. Only amino-acid replacements in the spike protein are shown, with predicted immune evasion mutations shown in bold (Table S1). Positive samples from bronchoalveolar lavage (BAL), endotracheal aspirates (ETA) or sputum are indicated in brown. Antibody-based anti-COVID19 treatments are represented by dashed vertical lines on the day of administration.

**Figure 4.**
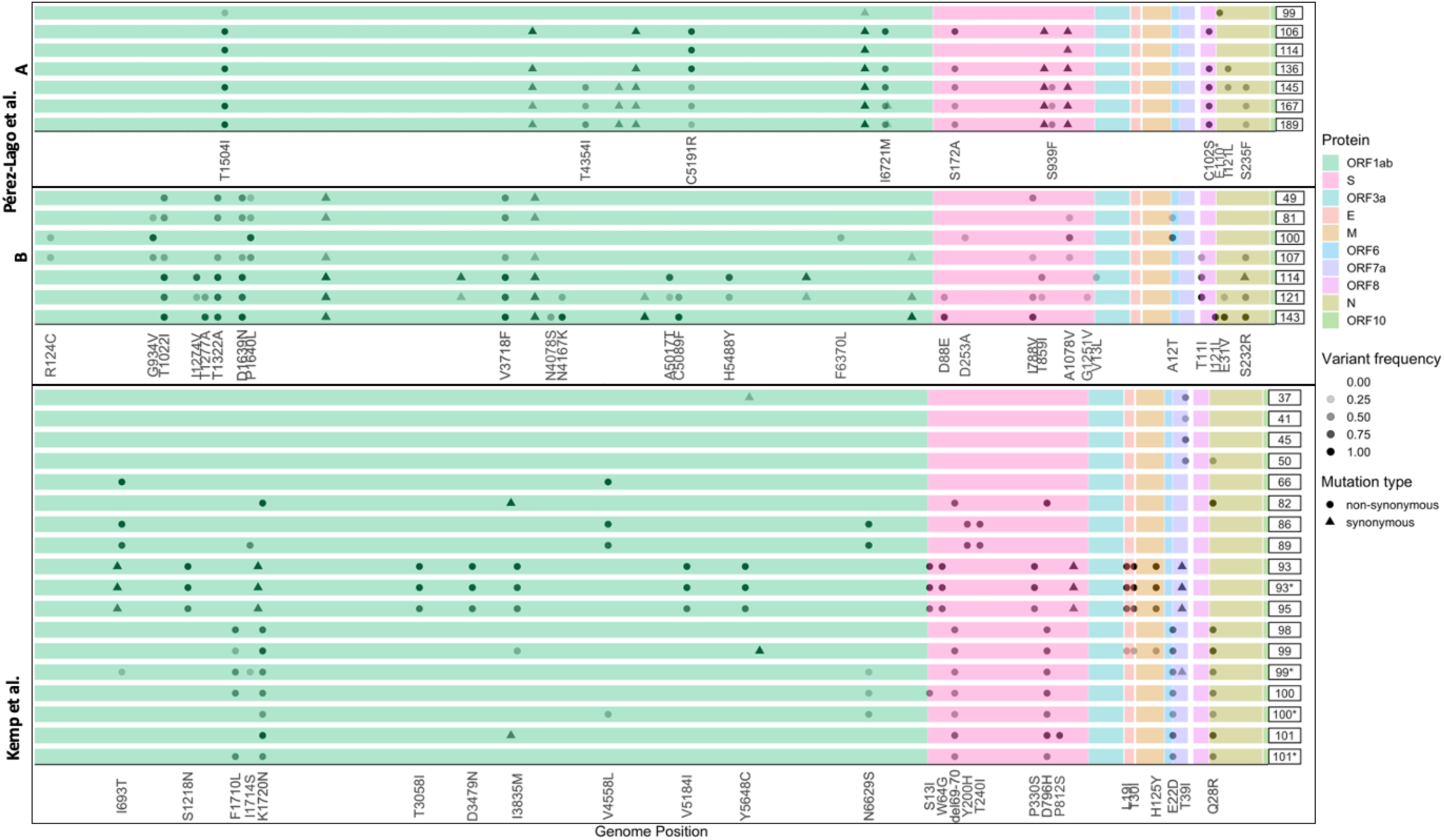
Polymorphic populations observed across patients. Each series of boxed lines represents a patient, and each line represents a sequenced time-point with time since infection on the right. The different open reading frames are color-coded. For each patient, only mutations relative to the first time point sequenced that appeared at a frequency ranging from 20% to 100% are shown.

## Predictors of antibody-evasion substitutions

We went on to examine the variation observed across different patients. When examining medical background, the patients could be roughly classified into one of the following categories: hematological cancers, HIV/AIDS, organ transplantation, and auto-immune disorders (Table 1). The latter two categories were often treated with steroids. Some but not all of the hematological cancer patients and others were treated with antibodies targeting B-cells, presumably causing profound B-cell depletion. In line with this, most of the patients with confirmed B-cell depletion showed negative serology for SARS-CoV-2 at one or more time-points (Table S1). Some patients were treated with ABT against SARS-CoV-2, whereas others were not, and in some antibody-evasion mutations were detected while in others they were not. Finally, we found that while in some ABT patients, immune evasion mutations were detected, sometimes these mutations fixed *before* the treatment (see e.g., Fig. 1, P5).

We noted that many patients (four of the six patients sequenced herein) displayed an intriguing cycling pattern of viral load (VL) (reflected by cycle threshold Ct values), with very high Ct values reaching negative or borderline negative results at one or more stages of the infection, followed by rebound of the virus. In the four abovementioned patients, this rebound was accompanied by clinical evidence for disease, highly suggestive of active viral replication. Several different hypotheses could explain this pattern. First, the virus may have cleared and been followed by re-infection with another variant. Since this pattern can be ruled out using sequencing, such cases were excluded from our analysis (Methods). Second, the virus may cycle between different niches, such as upper and lower airways. Its re-emergence in the upper airways (nasopharynx) may be due to selective forces or genetic drift. When considering selective forces, viral rebound may occur due to the near clearance of the virus, either via ABT or by the self-immune system, followed by the emergence of a more fit variant with antibody-evasion properties.

We set out to perform logistic regression to predict an outcome of antibody-evasion (Methods). We first tested if B-cell depletion or steroid treatment predict antibody-evasion mutations sometime during the course of infection (Table S2), and found this was not a significant predictor (P=0.6, P=0.4, N.S., respectively). Moreover, the length of sequenced infection (i.e., the maximal time-point sequenced) was not predictive of antibody-evasion either (P=0.9, N.S.). Next, we went on to test whether ABT predicts antibody-evasion, and if antibody-evasion tends to be preceded by a drop in viral load (Table S3). Our results showed that ABT is not a significant predictor (P=0.17, NS), whereas a drop in viral load followed by viral rebound is (P=2.6E-05) (Fig. 1). These results suggest that in some cases, what may be driving immune escape is actually the weakened immune system of the patient, although the somewhat low P-value of ABT suggests it is possible that the treatment itself also plays a role in driving antibody-evasion as well. This result suggests that the adaptive immune response of the patients herein is not completely absent, and that a weak antibody response is what drives adaptive evolution in the virus.

Next, we went on to examine patterns of variation over time across the different patients. In many of the case reports, the authors noted the emergence and disappearance (and sometimes re-emergence) of particular substitutions (Fig. 4). When re-analyzing the data, we noted that this pattern of variation across time was observed in the majority of patients (Table S2). From an evolutionary point of view, it is quite unlikely for one or more substitutions to disappear from a given population, and since we observe this at very different loci across all patients, it is not likely that this is due to recurrent sequencing problems or due to biases of the viral polymerase. We and others have previously noted sequencing errors that occur predominantly when viral load is low, when errors that occur during reverse transcription or early PCR cycles are carried over to higher frequencies ^10,11,41^. However, this phenomenon most often leads to errors in intra-host variants segregating at relatively low frequency, and is less common at the consensus sequence level, which is defined here as mutations present at a frequency of 80% or higher. We thus conclude that the most likely explanation for the highly dynamic polymorphisms observed, is the existence of distinct populations residing in different niches, which may correspond to different infected organs, or niches within an infected organ.

## Discussion

Tying the results obtained so far together, we suggest that viral rebound coupled with antibody-evasion may be enabled by partial penetration of antibodies to a specific niche. Accordingly, this partial penetration will prevent viral clearance on one hand, but on the other will promote selection for an antibody-evasion mutation. Notably, this is supported by the case of patient P1 in this study: during the time a negative nasopharyngeal swab was obtained, a BAL sample returned as positive, just prior to the emergence of the E484Q antibody evasion mutation. This suggests that the virus continued to thrive in lungs, possibly as a variant adapted to lungs, or as the same nasopharyngeal variant that migrated to the lungs. Alternatively, it is also possible that the immune evasion mutants continued to thrive in the nasopharyngeal swabs but were not picked up by the sample for PCR.

We go on to discuss the conditions that enable a variant in a chronic infections to keep spreading in the general population, and to potentially become a variant of concern. First, we searched for evidence of onwards transmission from a chronically infected patient in the cohort studied herein, yet found no such evidence in the publications or from epidemiological investigations of our TASMC patients. This is not necessarily surprising, as often such patients are bedridden, and in strict isolation both due to COVID-19 and due to their background conditions. However, this does raise the possibility that (a) chronic infections harbor variants that are non-infectious, and (b) chronic infections harbor non-transmissible virus, options we discuss below.

We first consider the option that chronic infections harbor non-infectious virus. In twelve of twenty-seven patients, evidence for viral replication was obtained, in the form of culturable virus, or in the form of PCR positive for sub-genomic RNA. In an additional eight patients, we noted either consistently high viral loads (Ct values around 20 or lower) or a negative difference of ten cycles between one PCR outcome and a later one (Table S6). In other words, in the latter scenario viral load increased dramatically across time in these patients, strongly suggestive of active viral replication. In line with this, we did not find evidence for mutations associated with defective virus, such as premature stop codons, frameshifts, or dramatic deletions outside of the spike protein (although in-frame deletions were noted in two patients, in ORF1a; Table S1). Thus, to summarize, while we cannot completely rule out the presence of non-infectious defective virus in chronic infections, we consider it is unlikely that all virus is non-infectious in most of the cases examined herein.

We next go on to discuss the probability that variants found in chronic infections are transmissible. In most of our patients (15/27) we observed either S:E484K/Q and/or deletions at S:Y144 in the NTD, both of which are highly recurrent mutations in the global phylogeny. However, globally circulating variants bearing these mutations were inferred to be less transmissible than other highly recurrent mutations (Table 2), and E484K/Q is notably absent in the once globally dominant variants Alpha and Delta. On the other hand, several mutations associated with higher transmissibility (e.g., S:P681R, ORF1a: Δ3675-3677) ^40^, were never observed in the chronic infections herein (Table 2).

Why if so were these mutations never observed? Evidently, in chronically infected patients we do not expect selection for mutations enhancing transmissibility, but rather expect selection to promote mutations that enhance viral replication (with immune evasion included in this latter category). However, there is often a link between better net viral replication and higher transmissibility ^42-44^, which should theoretically promote transmissibility-enhancing mutations also in in chronic infections. One possibility is that the sample size of patients in this study was too small. Given the large number of time we observed S:E484K/Q in chronic infections, it seems more likely that there is another explanation. Another possibility is that for example, S:P681H/R is under weaker positive selection in the conditions particular to immunocompromised patients. A recent study has suggested that part of the evolutionary advantage of S:P681H is its ability to evade the interferon response of the cell ^45^. Perhaps the particular conditions in immunocompromised prevent selection for interferon evasion: for example, some patients were treated with interferon inhibitors, although these are a minority of patients (Table S2); alternatively, the interferon response of immunocompromised patients is also likely weakened *per se*.

Finally, an additional possibility is that epistatic interactions among mutations prevent the emergence of some mutations, or in other words, two adaptive mutations may become deleterious when residing on the same genome. In line with this, the combination S: E484K/Q and S: P681H/R is not a common one, except for in Omicron (albeit E484A), where over 30 Spike amino-acid replacements have been observed. Indeed, several reports have suggested abundant epistatic interactions led to the emergence of Omicron ^46^, and these may allow the co-existence of S: E484A with S: P681H and others. It remains an enigma where Omicron first arose ^47^, yet the fact we see potentially epistatic interactions occurring in some chronic infections, coupled with the fact Omicron is characterized by many such pairs of mutations, suggest that lengthy chronic infections may allow an exploration of the SARS-CoV-2 fitness landscape, such that some of these chronic infections lead to the emergence of a highly transmissible variant.

## Conclusions

To summarize, we have found that the overall patterns of mutations observed in chronic infections closely mirror the pattern observed in VOCs, with some notable exceptions. We find that when viral rebound is observed, this strongly suggests that antibody-evasion mutations may have emerged in a COVID-19 patients. Thus, viral rebound can be viewed as a warning signal that VOC-like mutation occurred in a patient, and extra caution may be warranted: genetic sequencing, isolation, and close monitoring of contacts may be crucial when viral rebound is observed. While these immune-evasion mutations are of concern, we suggest that most variants emerging in chronically infected patients lack the potential for dramatic onwards transmission, possibly due to an absence of key mutations. Future research is necessary to understand the precise factors determining when and if a variant generated in chronic infection becomes highly transmissible. Overall, we suggest that extensive monitoring of chronic infections can be used for forecasting the set of mutations in future VOCs.

## Methods

### TASMC virus genome sequencing

Leftover RNA from nasopharyngeal swabs were obtained from six chronically infected individuals, across several different time points for each patients (Table S1). Patient 2 sequences were available from our previous study^16^; patients 1 and 3 were also previously sequenced ^15^, but here we added on additional time-points. All patients were positive via COVID-19 RT-qPCR tests of SARS-CoV-2 for a period longer than 3 weeks, and all were immunocompromised. All relevant clinical data was collected and was summarized in Tables 1, S2. All samples underwent whole-genome SARS-CoV-2 using the V3 ARTIC protocol (https://artic.network/ncov-2019) and the samples were sequenced using Illumina Miseq 250-cycle V2 kits at the Technion Genomic Center (Israel). This study was approved by the TASMC Helsinki committee (approval No. 1042-20-TLV), and by the Tel Aviv University IRB (approval number 0004435-1), with an exemption from informed consent.

### Determining genome consensus sequences and lineage determination

pTrimmer was used in order to trim the various primers that were used in the multiplex PCR (https://bmcbioinformatics.biomedcentral.com/articles/10.1186/s12859-019-2854-x). The raw reads were mapped to the reference genome of SARS-CoV-2 (GenBank ID <u>MN908947</u>). Mapping and variant calling was performed using our AccuNGS pipeline ^41^. The consensus sequence of each sample was determined based on sites with coverage of at least 10X sites. As we and other have observed, the sequencing process may lead to erroneous mutations being observed, and this is particularly exacerbated at low viral loads ^10,11,41^. To this end, only mutations with a frequency of at least 80% were introduced into the consensus sequence (hereby referred to as substitutions or fixed mutations); loci with mixed populations at frequencies lower than 80% were considered ambiguous and marked by “N”.

For each patient, substitutions as compared to the reference that appeared in the first time point were marked as background substitutions, and were not considered for any further analyses. Only substitutions that were added in the following time points were considered. The “Pangolin” network was used to identify the consensus sequence lineage (https://cov-lineages.org/resources/pangolin.html). This allowed us to rule out re-infection in all patients but one: the last time-point of P6 from day 48 of infection was inferred to be due to re-infection (B.1.1.7 as compared to B.1.1.50 in all earlier time-points). The last time point from this patients was removed from the analysis, leaving a total of 37 days. Visual inspection of the results led to a suspected contamination at day 27 of P5, as several B.1.1.7 lineage defining mutations were observed on an otherwise B.1.1.50 background. This sample was omitted from the analysis.

### Data availability

All consensus sequences for the patients sequenced herein were deposited in GISAID, and raw sequencing reads were uploaded to the NCBI Sequence Read Archive under submission number PRJNA803960 (Table S5).

### Reanalysis of previous case reports

Previously published case reports were selected by mining the literature for the key-words “prolonged”, “persistent” or “chronic” “SARS-CoV-2 infection” up till Nov. 15 2021. While some studies validated the chronic nature of infection by performing culturing of the virus or by measuring sub-genomic RNA (sgRNA), several did not. Thus, when culture/sgRNA data was unavailable, we filtered out patients with infections of less than 20 days, based on an a meta-analysis that showed viral shedding was mostly limited to 20 days (see main text). We verified intermediate to high viral load (Ct values of 27 or lower) at some time exceeding day 20 as a proxy for ensuring that shedding was not of residual non-infectious virus. Only studies that performed longitudinal whole genome sequencing were maintained. In Tarhini. et al.^25^ patient 3 was removed from the analysis as the authors identified a co-infection event. Sequencing data for Jensen et al.^22^ was obtained from the authors, albeit with missing data – the first time point for patient D was missing. We thus relied on the report by the authors on S:E484 at time point 1, and only mutation S:E484K was considered in time point 2 for patient D.

When analyzing the data, and in line with the above, the focus of almost all our analyses, was on mutations with a frequency higher than 80%, not present in the first time point sequenced. When analyzing viral polymorphisms across time (Fig. 4) we also present mutations at lower frequencies. All relevant clinical data was collected and was summarized in Tables 1 and S2. Ct values for RT-qPCR tests were also extracted from all publications (Table S6). A test was considered borderline if the Ct ranged from 34 to 37, and was considered negative if reported as such, or was equal to 38 or higher. In one study ^22^, raw viral load measurements were presented in terms of number of copies per ml sample, and a negative result was reported when zero copies were detected.

### Summary information of the patients

All available medical records including the patient’s background condition, treatments and anti SRAS-CoV-2 serology test are specified in Table S2. Our cohort composed of many immunocompromised conditions: seven-teen of the patients suffer from different kinds of lymphomas and leukemias, four of the patients undergo organ transplant, tree suffer from AIDS, two of the patients have an autoimmune disease and one suffer from renal disease. Regarding age groups: ten of the patients are classified in age range of 60-69, five belong to 70-79, and tree patients to 40-49 years old. In each of the age ranges of 0-9, 20-29, 30-39 and 50-59 two patients were added and only one patient is in age range of 80-89 years old.

### Summary of substitution events

A summary of all substitution events was compiled into three tables. Table S1 lists every substitution and the patient/s where it was found. Data on the proportion of a given substitution in the general global viral samples was taken from http://cov-glue-viz.cvr.gla.ac.uk/, and counts of substitutions along a sampled phylogeny of approximately 3,500 sequences were taken from NextStrain (www.nextrain.org) ^48^ accessed on Jan 2, 2022. Each of the spike amino-acid replacements was queried to see if information exists on antibody-evasion for a given mutation, and was marked as enhancing antibody-evasion if a publication was available showing direct experimental evidence supporting this (Table S1).

Table S2 lists information (clinical and mutational, across all time-points) per patient, whereas Table S3 lists information per time point sequenced for each patient. When two samples from the same time point were available from different niches, we presented only the nasopharyngeal one, since our focus is primarily on this niche. Counts and information of substitutions per patient are listed in table S2 for both the spike region and the rest of the genome. Transient substitutions (that re-appeared at a certain time point) were counted only once.

### Substitution events histogram

We counted the total number of substitutions observed at each position in the genome in three sets of sequences:

a. our set of chronic infections
b. the lineage defining mutation of the five VOCs (Alpha, Beta, Gamma, Beta, Omicron), as defined by https://covariants.org. Substitutions shared by two or more VOCs were omitted.
c. a subset of sequences from NextStrain (www.nextrain.org)^48^ accessed on Jan 2, 2022. Sequences were filtered to only those earlier than 16/8/2020. The numbers of events per genome position were downloaded from the site.

A histogram was created for each of the three sets, with bin sizes of 500 bases. The probability of observing a given number of substitutions in a bin was calculated based on a binomial distribution with p=500/L (L being the reference genome length), n=the sum of all substitutions observed in a set, and k=observed number of substitutions in a given bin. Correction for multiple testing was performed using the false discovery rate (FDR) of Benjamini–Hochberg^49^.

### Logistic regression

Logistic regression was performed twice to test for significant predictors of an antibody-evasion outcome. First, at the level of each patient, we tested whether severe B-cell depletion, steroid treatment, or the maximal time-point since infection that was sequenced. Severe B-cell depletion was defined as one of the following conditions: treatment with anti-B cell directed antibodies up to nine months prior to COVID-9 detection, or blood-test measurements of low B-cell markers. Second, at the level of time-point sequenced, we tested whether ABT (convalescent plasma, hyperimmune plasma, or monoclonal antibody/ies) or viral rebound predicted an outcome of an antibody-evasion mutation. Each sequenced time point was hence characterized as ABT-true if treatment was given up to seven days before the treatment, and viral rebound was defined as true if the COVID-19 PCR test just prior to the sequenced time point was borderline or negative. Binomial logistic regression was performed using the glm package in R. All the raw data is available in tables S2 and S3.

### Co-occurring substitutions

We began by identifying all pairs of substitutions that occurred in two or more patients. A *χ*^2^ test was used in order to identify a deviation from expected frequencies, by comparing marginal probabilities with paired probabilities (number of degrees of freedom was 1). Multiple testing was accounted for via FDR ^49^.

## Data Availability

All consensus sequences for the patients sequenced herein were deposited in GISAID, and raw sequencing reads were uploaded to the NCBI Sequence Read Archive under submission number PRJNA803960. Full details are available in the manuscript supplementary material.

## Acknowledgements

We would like to thank Shay Fleishon, Neta Zuckerman, Tzachi Hagai, David Burstein, and Noam Harel for critical reading and discussions. This study was supported by an ERC starting grant 852223 (RNAVirFitness), and by an Israeli Science Foundation grant 3963/19. This study was supported in part by a fellowship to SH from the Edmond J. Safra Center for Bioinformatics at Tel-Aviv University. The funders had no role in study design, data collection and analysis, decision to publish or preparation of the manuscript.

